## Supplementary figures 1-9 and Supplementary Table 1 for "Immunosuppression causes dynamic changes in expression QTLs in psoriatic skin"

### Supplementary Information

#### Supplementary Figures

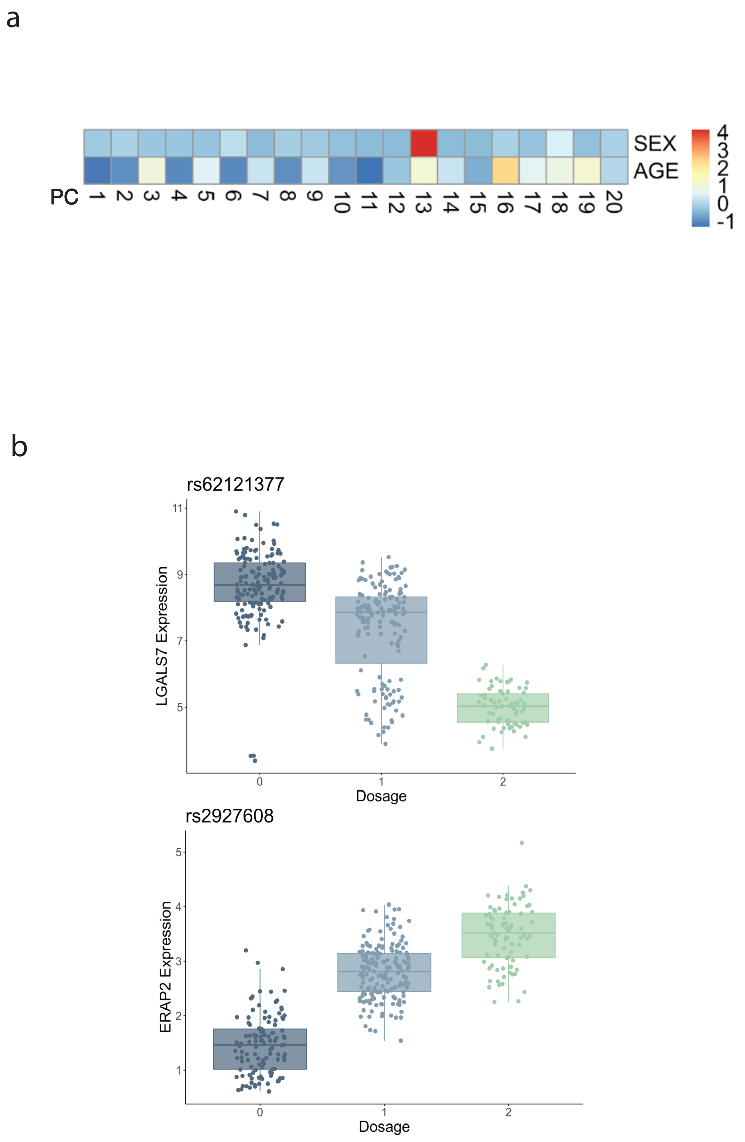


**Supplementary Figure 1. Mapping eQTLs with PAUSE trial skin biopsies. (a)** To account for potential confounding effect by gene expression variation, 20 RNAseq PCs were included as covariates in the model. Clinical characteristics such as sex and age were not included, as they were captured in the 20 PCs. **(b)** Expression level of genes *LGALS7* and *ERAP2* plotted with respect to rs62121377 and rs2927608 genotypes.

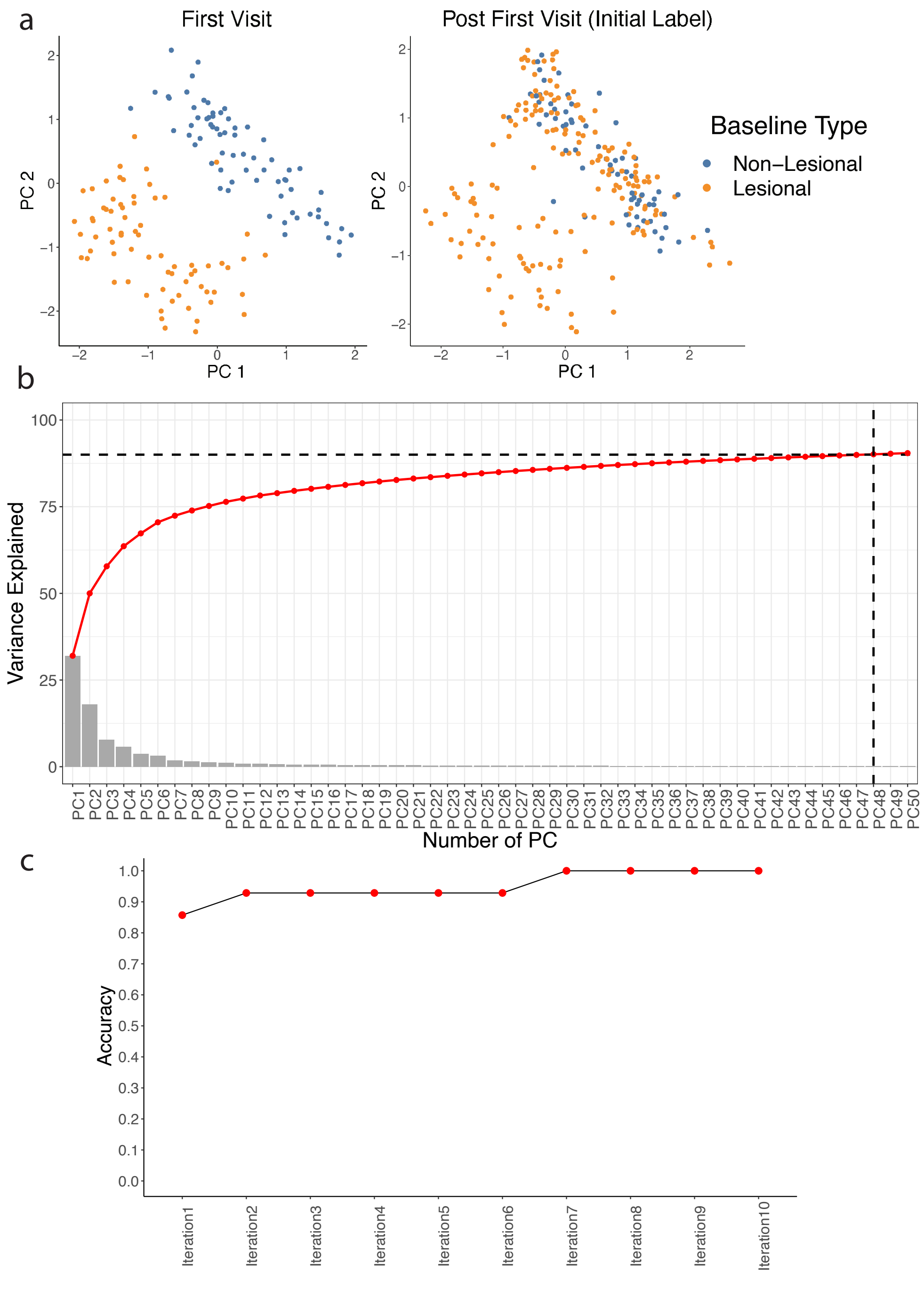


**Supplementary Figure 2. Skin biopsy transcriptome in PCA space. (a)** All the samples were labelled as lesional or non-lesional based on the skin status at the first visit. The first 2 RNAseq PCs illustrate the shift of transcriptional profile of lesional samples to a more non-lesional like profile as the lesion resolved over the study. **(b)** The first 48 PCs explain >90% of the variance of the transcriptional data. **(c)** Performance of LDA classifier using SPITS score to predict inflammation status in 10-fold cross-validation**.**

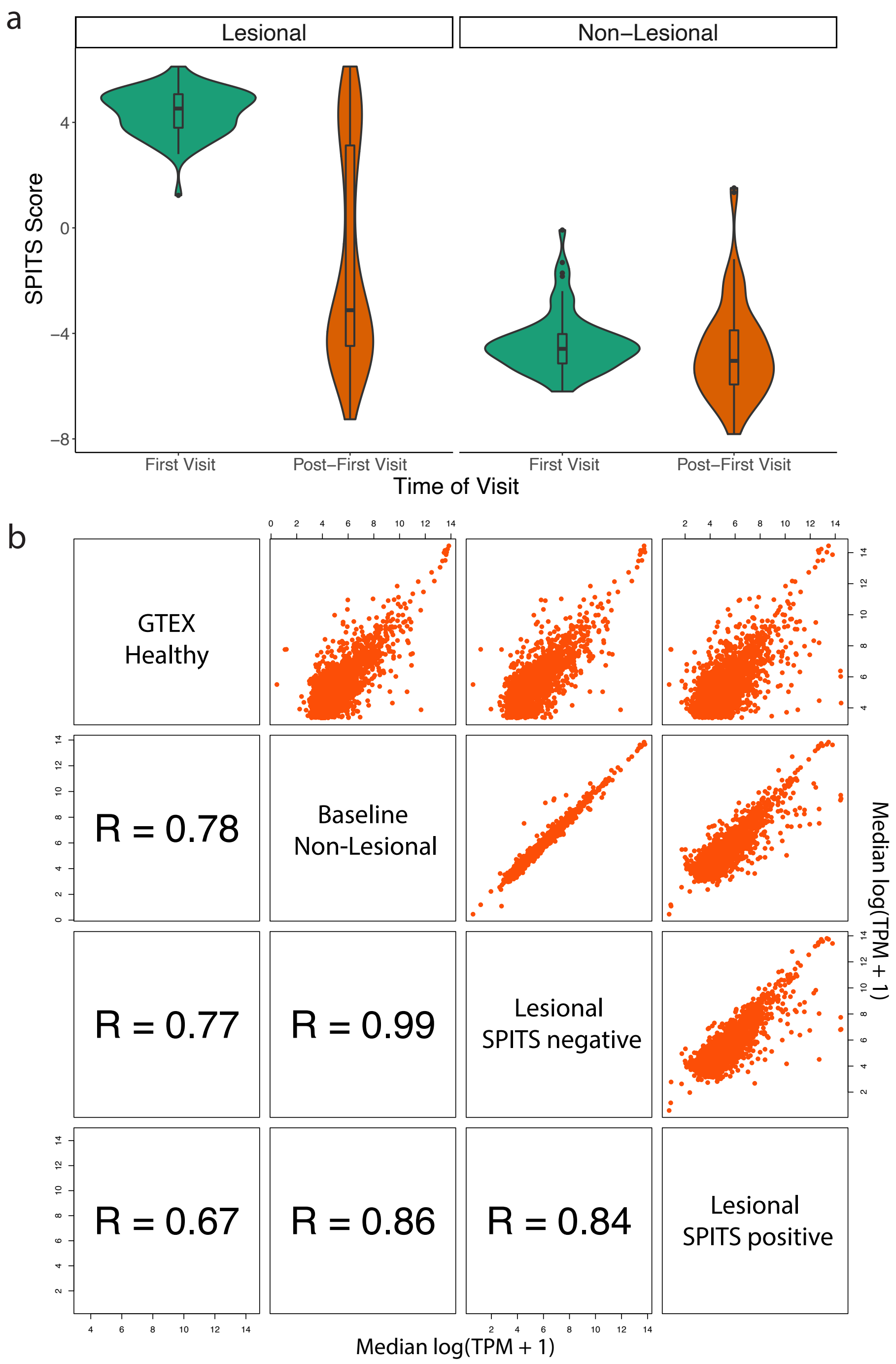


**Supplementary Figure 3. SPITS distribution and correlation across samples. (a)** SPITS score distribution in lesional and non-lesional samples at the first visit and after the first visit. **(b)** Scatterplot matrix of gene expression from healthy samples, baseline non-lesional samples, SPITS negative lesional samples and SPITS positive lesional samples. Healthy skin samples from GTEx consortium[1] are more correlated with baseline non-lesional and SPITS negative lesional samples comparing to SPITS positive lesional samples.

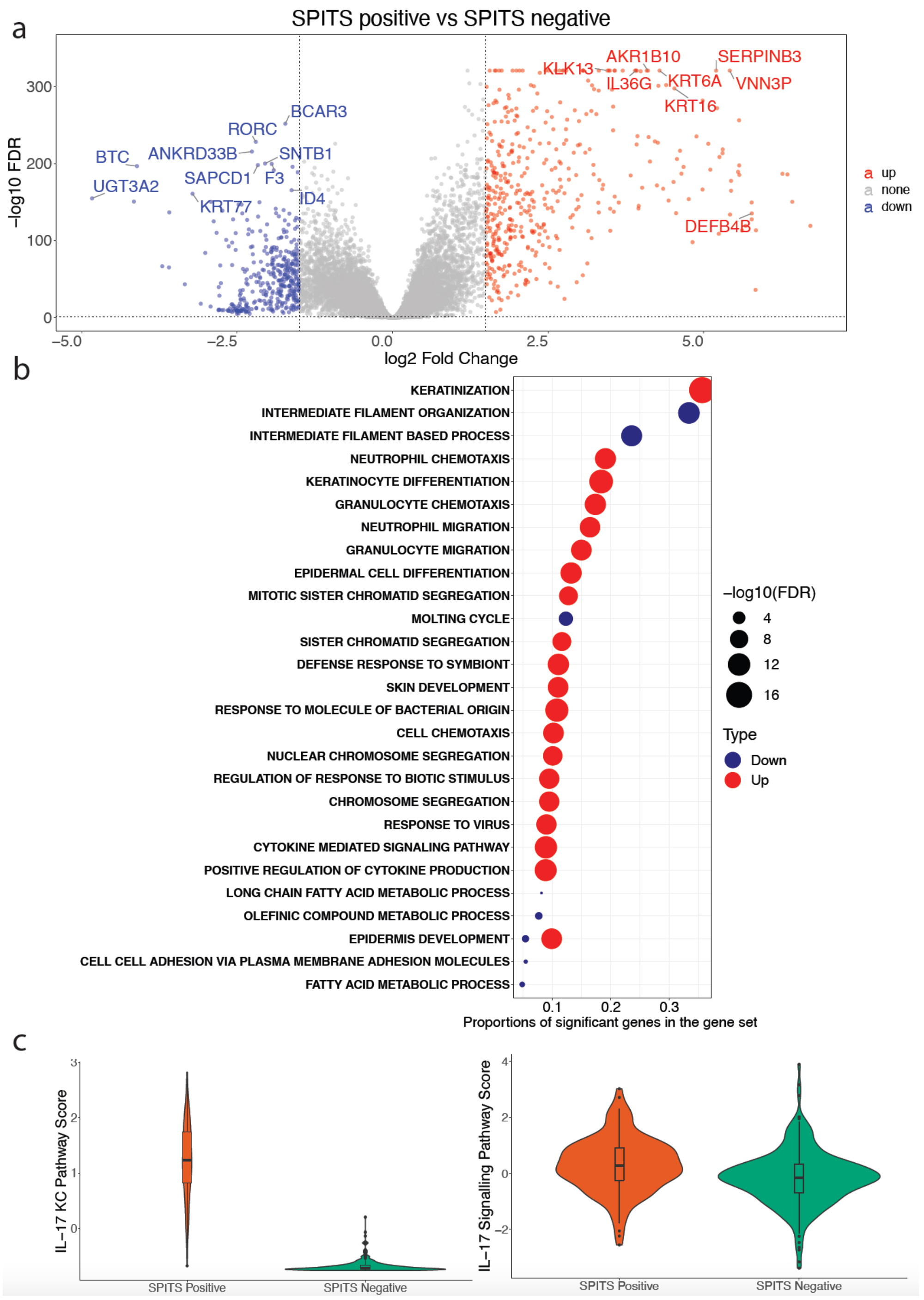


**Supplementary Figure 4. Biologocal interpretation of SPITS. (a)** Differentially expressed genes(DEGs) between SPITS positive and SPITS negative samples. Differential expression is defined by FDR < 0.05 and |logFC| > 1.5. Upregulated genes are colored in red, downregulated genes are colored in blue, and non-significant genes are colored in grey. **(b)** GO enrichement analysis of DEGs**. (c)** *IL-17* pathway scores generated from induced keratinocytes[2] in SPITS positive and SPITS negative samples (left), *IL17* pathway scores generated from curated gene set[3] in SPITS positive and SPITS negative samples (right).

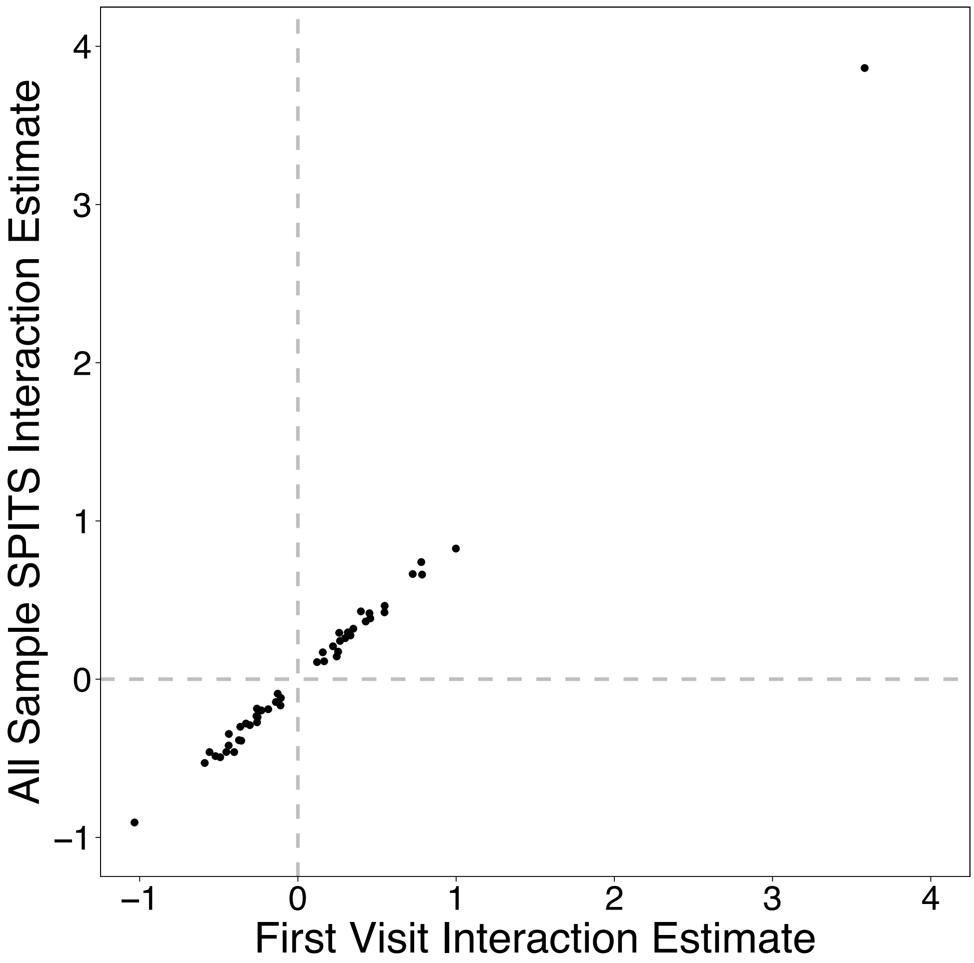


**Supplementary Figure 5. Concordance of inflammation-eQTL interaction effects.** eQTL interactions observed using only first visit samples (i.e. baseline biopsy type-eQTL interaction) are plotted against interactions observed with all samples (i.e. SPITS-eQTL interaction). Each point represents a significant interaction (FDR < 0.20) present in both scenarios (N = 49). The estimates are strongly correlated (R = 0.99, p < 1e-26)

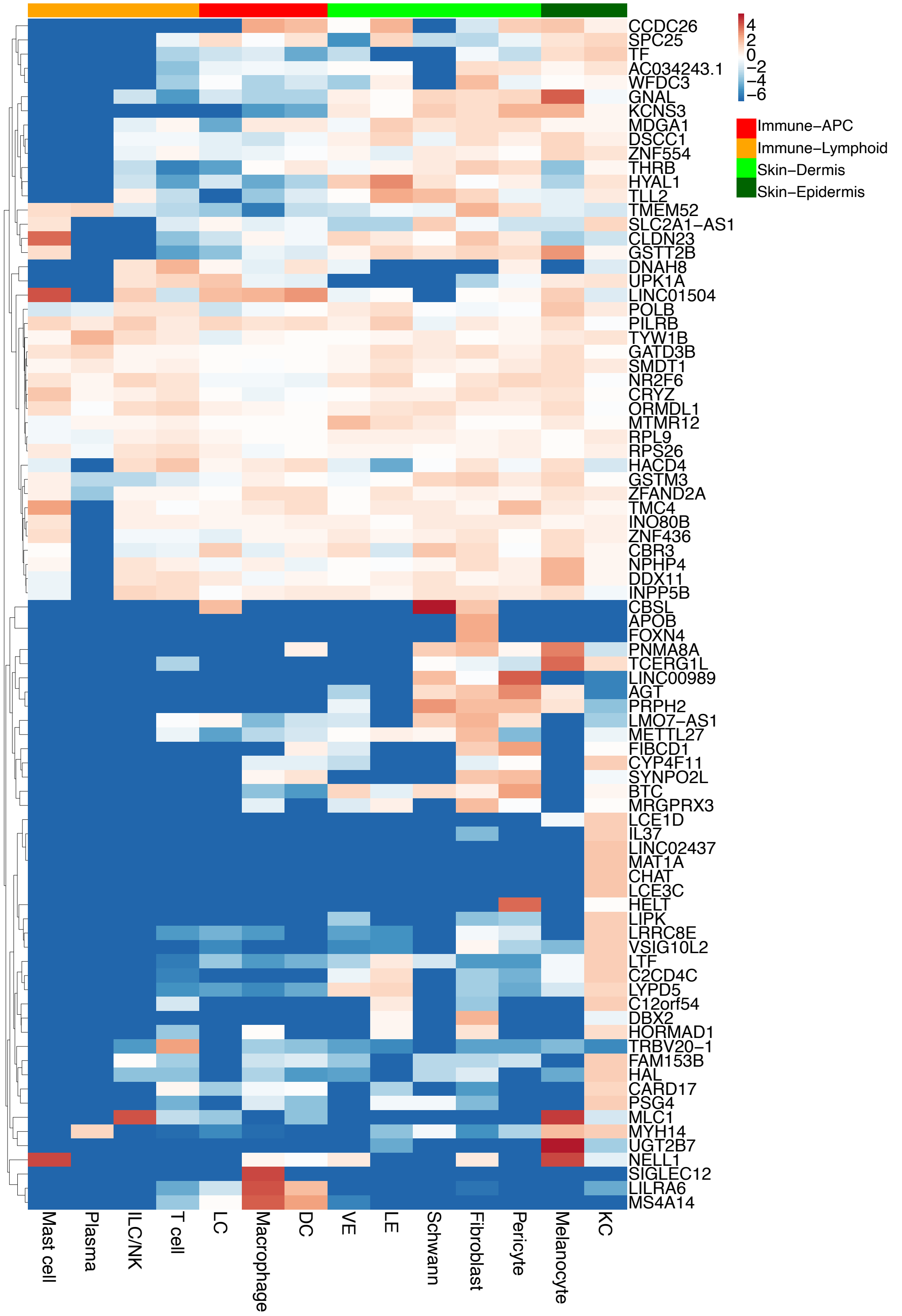


**Supplementary Figure 6. Log fold change of interacting eGene expression per cell type against mean eGene expression across the 14 cell types.** The cell types are classified into 4 categories: immune-antigen presenting cells (APC), immune-lymphoid, skin-epidermis and skin-dermis.

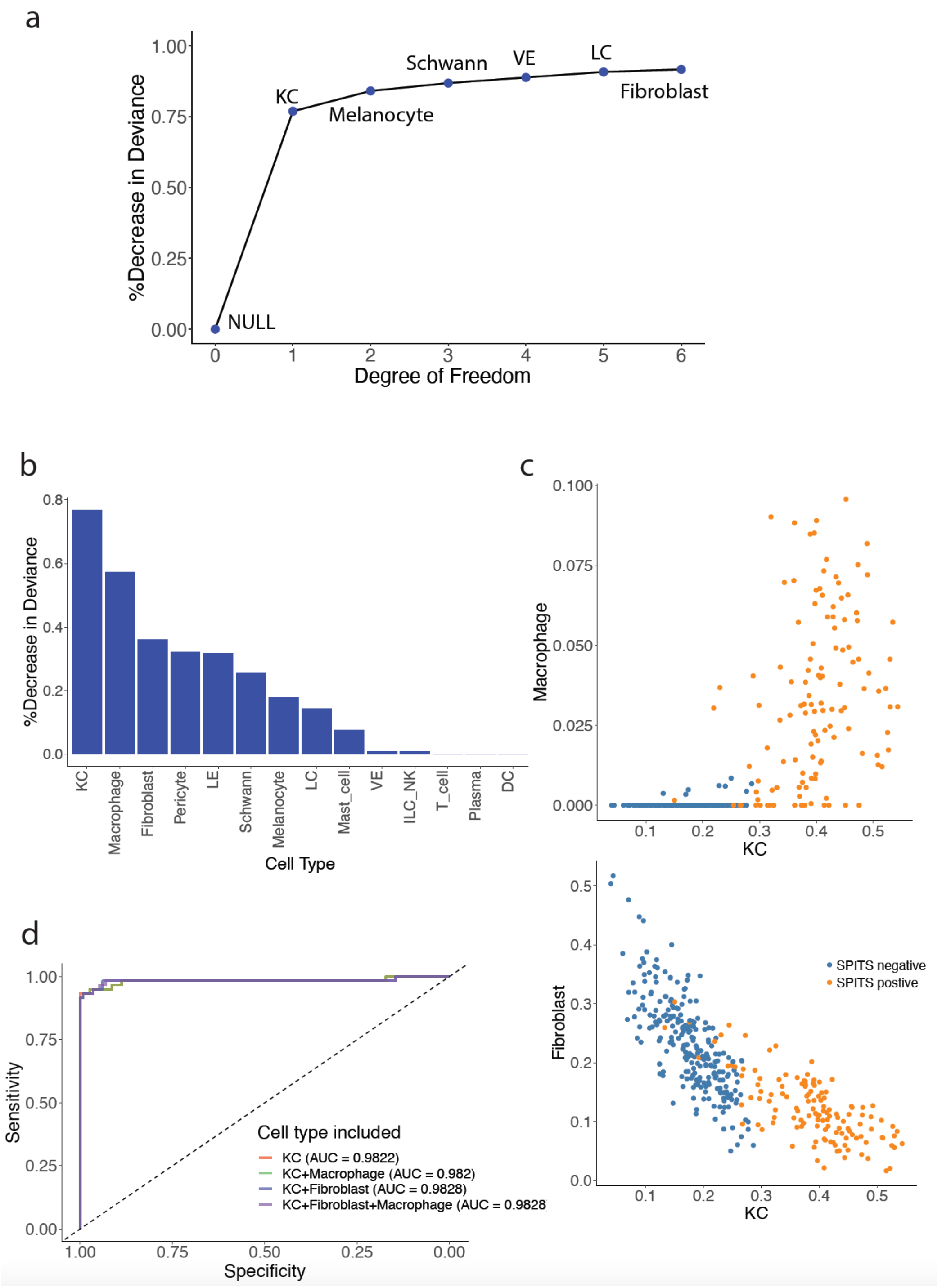


**Supplementary Figure 7. Cell type composition predicts SPITS. (a)** %Decrease in deviance for the selected logistic model. KC, keratinocyte; VE, vascular endothelium; LC, Langerhans cells. **(b)** Decrease in deviance for univariate models. **(c)** Keratinocyte (KC), macrophage and fibroblast fractions separate inferred inflammation status. **(d)** ROC curves evaluating LDA classifiers using different cell types as predictors.

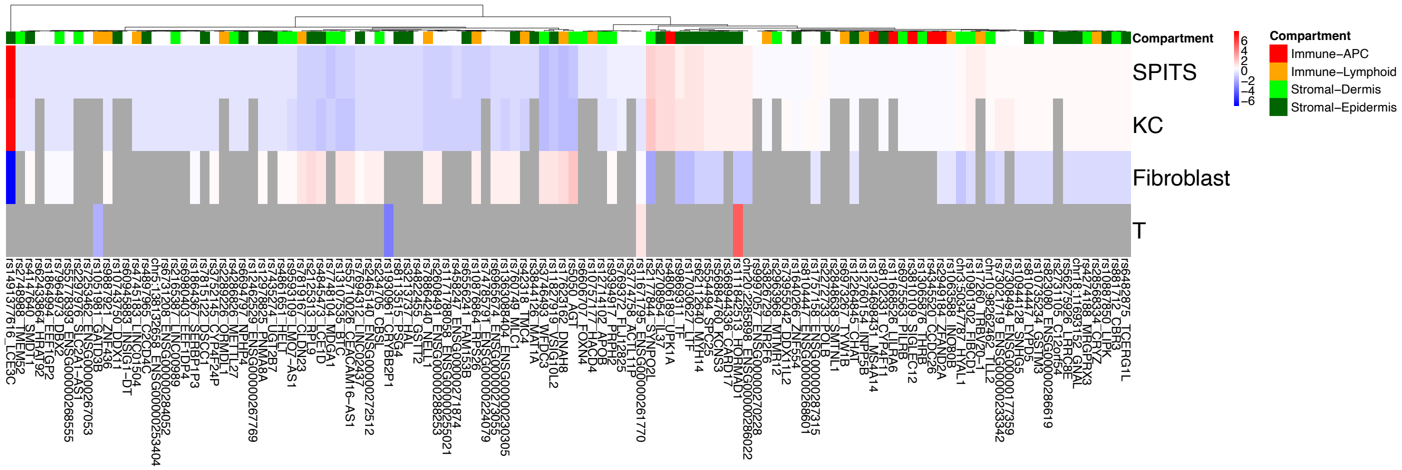


**Supplementary Figure 8. Heatmap of all SPITS eQTL interactions, in comparison with KC, fibroblast, and T cell interactions.** The values are the absolute value of interaction coefficient z-scores. The dampeners are colored in blue, the magnifiers colored in red, and non-significant interactions are in grey. Tissue specificity of the interactions are annotated using the same eGene tissue-specificity identified with single-cell data. No tissue specificity labeled if the eGene was not present in the single-cell data.

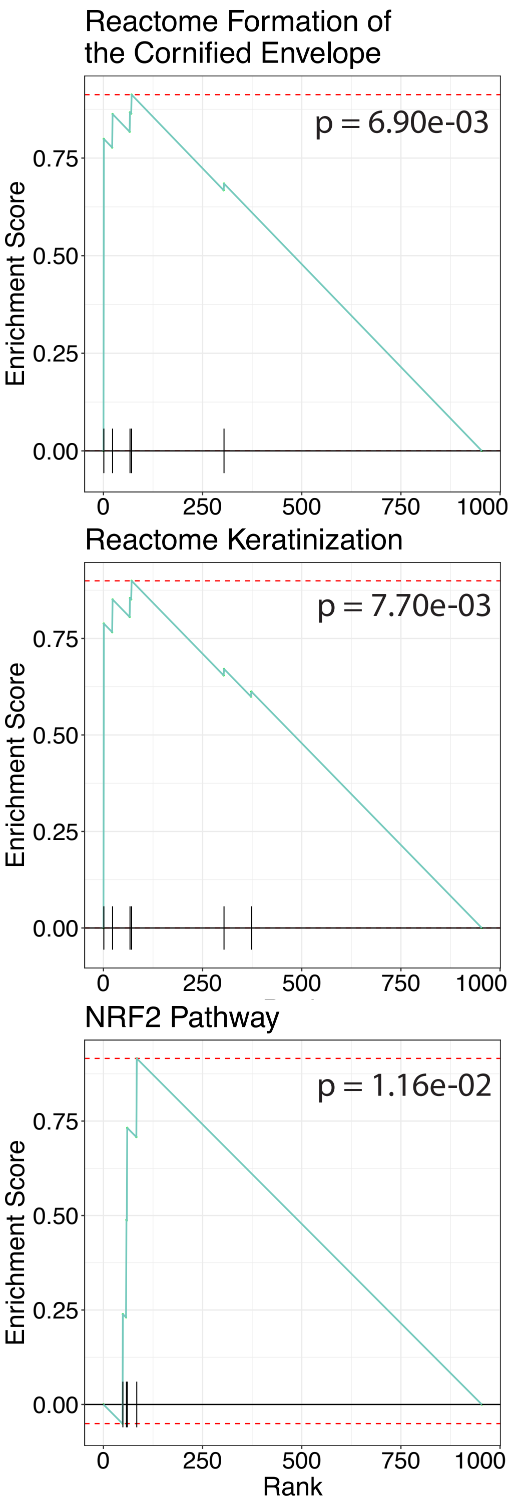


**Supplementary Figure 9. Gene set enrichment analysis of SPITS-interaction eGenes.** The genes are ranked by absolute value of interaction beta.

#### Supplementary Table

**Supplementary Table 1. PAUSE Trial[4] Participant Demographic and Clinical Characteristics.** BMI, body mass index; PASI, psoriasis and area severity index.

| | **Number (%) of Participants** | |
|---|---|---|
| **Characteristic** | **Total (n = 108)** | **Included for eQTL analysis (n = 77)** |
| Age, mean (SD), y | 46.1 (12.08) | 47.2 (11.8) |
| Sex | | |
| Male | 73 (67.6) | 51 (66.2) |
| Female | 35 (32.4) | 26 (33.8) |
| Race | | |
| White or Caucasian | 95 (88.0) | 64 (83.1) |
| Black or African American | 6 (5.6) | 6 (7.8) |
| Asian | 4 (3.7) | 4 (5.2) |
| Other | 1 (0.9) | 2 (2.6) |
| Multiple Races | 2 (1.9) | 1 (1.3) |
| Ethnicity | | |
| Hispanic or Latino | 12 (11.1) | 6 (7.8) |
| Not Hispanic or Latino | 96 (88.9) | 71 (92.2) |
| Weight, mean (SD), kg | 97.5 (21.48) | 96.7 (21.68) |
| Height, mean (SD), cm | 171.9 (9.87) | 171.5 (10.19) |
| BMI, mean (SD), $kg/m^2$ | 33.0 (6.88) | 32.9 (7.21) |
| Week 0 PASI | | |
| Number | 108 | 74 |
| Mean (SD) | 19.9 (8.06) | 19.9 (8.50) |
| 12-20 | 71 (65.7) | 49 (67.6) |
| >20 | 37 (34.3) | 24 (32.4) |
| Week 12 PASI | | |
| Number | 103 | 70 |
| Mean (SD) | 3.1 (4.27) | 2.1 (1.99) |
